# Plasma Neurofilament Light Chain predicts cognitive progression in prodromal and clinical dementia with Lewy Bodies

**DOI:** 10.1101/2021.03.19.21253993

**Authors:** Andrea Pilotto, Alberto Imarisio, Claudia Carrarini, Mirella Russo, Stefano Masciocchi, Stefano Gipponi, Elisabetta Cottini, Dag Aarsland, Henrik Zetterberg, Nicholas J. Ashton, Abdul Hye, Laura Bonanni, Alessandro Padovani

**Affiliations:** Neurology Unit, Department of Clinical and Experimental Sciences, University of Brescia, Brescia, Italy; FERB Onlus, Ospedale S. Isidoro, Trescore Balneario, Bergamo, Italy; Department of Neuroscience Imaging and Clinical Sciences, University G. d’Annunzio of Chieti-Pescara, Chieti, Italy; Division of Pharmacology, Department of molecular and Translational Medicine, University of Brescia, Brescia, Italy; Centre for Age-Related Medicine, Stavanger University Hospital, Stavanger, Norway and Department of Old Age Psychiatry, Institute of Psychiatry, Psychology & Neuroscience, Maurice Wohl Clinical Neuroscience Institute, King’s College London, London, UK; Department of Psychiatry and Neurochemistry, Institute of Neuroscience and Physiology, The Sahlgrenska Academy, University of Gothenburg, Gothenburg, Sweden; Clinical Neurochemistry Laboratory, Sahlgrenska University Hospital, Mölndal, Sweden; Department of Neurodegenerative Disease, UCL Institute of Neurology, London, UK; UK Dementia Research Institute at UCL, London, UK; Wallenberg Centre for Molecular and Translational Medicine, Department of Psychiatry and Neurochemistry, Institute of Neuroscience and Physiology, the Sahlgrenska Academy at the University of Gothenburg, Sweden; King’s College London, Institute of Psychiatry, Psychology & Neuroscience, Maurice Wohl Clinical Neuroscience Institute, London, UK; NIHR Biomedical Research Centre for Mental Health & Biomedical Research Unit for Dementia at South London & Maudsley NHS Foundation, London, UK

**Keywords:** Dementia with Lewy Bodies, neurofilament light chain, cognitive progression, biomarkers

## Abstract

Plasma neurofilament light chain (NfL) is a marker of neuronal damage in different neurological disorders and might predict disease progression in dementia with Lewy bodies (DLB). The study enrolled 45 controls and 44 DLB patients (including 17 prodromal cases) who underwent an extensive assessment at baseline and at 2 years follow-up. At baseline, plasma NfL levels were higher in both probable DLB and prodromal cases compared to controls. Plasma NfL emerged as the best predictor of cognitive decline compared to age, sex and baseline severity variables. The study supports the role of plasma NfL as a useful prognostic biomarker from the early stages of DLB.

## INTRODUCTION

Dementia with Lewy Bodies (DLB) is a complex neurodegenerative disorder characterized pathologically by misfolding and aggregation of alpha-synuclein, as well as A-beta and tau proteins, and by a combination of cognitive deficits with fluctuations, visual hallucinations, parkinsonism, REM-sleep behavior disorder (RBD) which represent the core clinical symptoms and is often associated with behavioral abnormalities and autonomic symptoms [1–4]. The prognosis is poor [5] and the variation in cognitive decline is large [6].

Despite the recent advances in the clinical definition of DLB from prodromal stages, disentangling the heterogeneity of presentation and disease progression of patients is still a great challenge for the research community [3].

Diagnosing and stratifying DLB patients are pivotal for a proper personalization of management strategies and identifying patients with different rate of progression will enable a proper selection of patients in incoming disease-modifying interventions.

Neurofilament light chain (NfL), a highly expressed protein in large caliber myelinated axons, has been recently proposed as general marker of neuronal degeneration and damage in different neurological disorders [7,8]. In fact, CSF and plasma NfL levels are significantly altered in Alzheimer’s disease (AD), prion diseases, amyotrophic lateral sclerosis, frontotemporal dementia, whereas findings in Parkinson’s disease (PD) and early DLB were contradictory [9]. In addition to that, it has been reported that plasma and CSF NfL levels correlated with disease severity and progression in PD [8,10,11] and Alzheimer’s disease [12]. Very few studies have focused on DLB patients and, to the best of our knowledge, no prospective longitudinal studies evaluated plasma NfL as potential early diagnostic and prognostic marker in DLB. Therefore, the aim of the present study was to investigate the relationships between plasma NfL and motor and cognitive symptoms in DLB from the prodromal stage and to evaluate its accuracy as predictor of progression after two-years of follow-up.

## METHODS

### Patients selection and assessment

Consecutive patients with a clinical diagnosis of probable DLB [2] or prodromal DLB [3] were enrolled between October 2016 and March 2018 for the baseline evaluation at the Neurology Unit at the University of Brescia, Italy and the Neurology Unit at the University G. d’Annunzio of Chieti-Pescara, Chieti, Italy. Healthy controls subjects (HC) with normal cognitive and motor functions were recruited among patients’ caregivers. This study was approved by the local ethics committee (NP 1471, DMA, Brescia) and was in conformity with the Helsinki Declaration; informed consent was obtained from all participants. All patients underwent routine blood analyses, magnetic resonance imaging (MRI) and [^123^I]FP-CIT SPECT imaging supporting the diagnosis [13]; levodopa equivalent daily dose (LEDD) was calculated according to standard conversion [14] and the diagnosis was supported by at least one year of clinical follow-up.

The following exclusion criteria were applied: (1) cognitive deficits or dementia due to other causes than DLB; (2) prominent cortical or subcortical infarcts or brain/iron accumulation at imaging; (3) other neurologic disorders or medical conditions potentially associated with cognitive deficits; (4) bipolar disorder, schizophrenia, history of drug or alcohol abuse or impulse control disorder; (5) recent traumatic events or acute fever/inflammation (potentially influencing NfL levels).

At baseline, standardized neurological examination was performed, including the Movement Disorder Society-Unified Parkinson Disease Rating Scale, part III (MDS-UPDRS, part III) [15], a full neuropsychological assessment including the Mini-Mental State Examination (MMSE), the Neuropsychiatric Inventory (NPI) for the assessment of global cognitive and psychiatric functions [16] and an evaluation of basic and instrumental activities of daily living [17]. DLB patients without dementia (i.e., independent in activities of daily living) at baseline and two years - follow-up were retrospectively re-classified as prodromal DLB according to recently proposed diagnostic criteria [3].

All patients included in the analyses underwent a clinical follow-up at 2 years including full neurological examination, motor, cognitive and behavioural assessment.

### Biochemical analyses

At the time of assessment, approximately 10 mL venous blood was collected in glass tubes containing sodium ethylenediaminetetraacetic acid (EDTA) from each subject. The blood samples were centrifuged at 2000 x g at 4 °C for 8 min within 2 h of collection. Plasma supernatant was collected, divided into aliquots, and frozen at −80 °C until further use. Plasma NfL concentration was measured using the Simoa HD-1 platform (NF-Light; Quanterix, Billerica, MA) at the Maurice Wohl Clinical Neuroscience Institute, London, UK. Samples were randomized, blinded and measured in duplicate using a batch of reagents from the same lot. The intra-assay and inter-assay coefficients of variation were 8.1 and 11.2%, respectively. The lower limit of quantification was 0.136 pg/mL and the lower limit of quantification (LLOQ) was 0.696 pg/mL when compensated for a 4-fold sample dilution. Outliers with plasma NfL value above more than 5 standard deviations of the mean were excluded from the study analysis (n = 1 probable DLB; n = 1 HC) [18].

### Statistical analyses

Group differences between HC and total DLB, as well as between probable DLB and prodromal DLB were assessed with Mann-Whitney U test or Fisher exact test for continuous or dichotomous variables, respectively. Plasma NfL levels between groups were compared with age-adjusted one-way ANOVA. Progression of MMSE, MDS-UPDRS-III and NPI between groups was evaluated with repeated measures ANOVA adjusted for age, sex, disease duration, baseline MMSE and MDS-UPDRS-III (for MMSE and MDS-UPDRS-III progression) and for age, sex, disease duration, baseline MMSE and baseline NPI (for NPI progression).

Plasma NfL levels and established prognostic factors (age, gender, disease duration, MDS-UPDRS III score, MMSE and NPI) were analyzed by a multivariable linear regression in order to evaluate the best predictors of changes within cognitive (MMSE), behavioral (NPI) and motor (MDS-UPDRS-III) scores at two years. Significance was set at p < 0.05 for all the analysis. SPSS 24 (IBM, Armonk, NY) was used for statistical analyses.

## RESULTS

### Recruitment, clinical and cognitive baseline features

Forty-four DLB patients (including 27 with dementia and 17 prodromal cases) and forty-five healthy controls were consecutively enrolled. Three patients deceased during the follow-up and 41 patients were thus included in the longitudinal analyses. Between prodromal DLB patients (n = 17), n = 13 were diagnosed as probable MCI-LB, n = 3 as psychiatric-onset prodromal DLB and n = 1 as delirium-onset prodromal DLB. Clinical and demographic features of DLB with dementia and prodromal cases are shown in Table 1. According to the definition, prodromal cases were independent for activities of daily living and exhibited higher total MMSE scores compared to DLB with dementia (25.5 ± 1.3 vs 18.3 ± 3.7; p = 0.001). HC were slightly younger than DLB patients (73.3 ± 7.6 vs 77.5 ± 5.4 years respectively, p = 0.003) and comparable for sex distribution (44.4% vs 40.9% males respectively, p = 0.736). In age-adjusted comparisons, NfL levels were higher in DLB total cohort and separately in both probable DLB and prodromal cases compared to HC (Figure 1 and Table 1, HC 25.7 ± 10.5 pg/ml, DLB with dementia 55.3 ± 20.9 pg/ml, prodromal DLB 46.9 ± 23.2 pg/ml; total DLB vs HC: p < 0.001; prod-DLB vs HC: p = 0.001; probable DLB vs HC: p < 0.001).

**Table 1.**
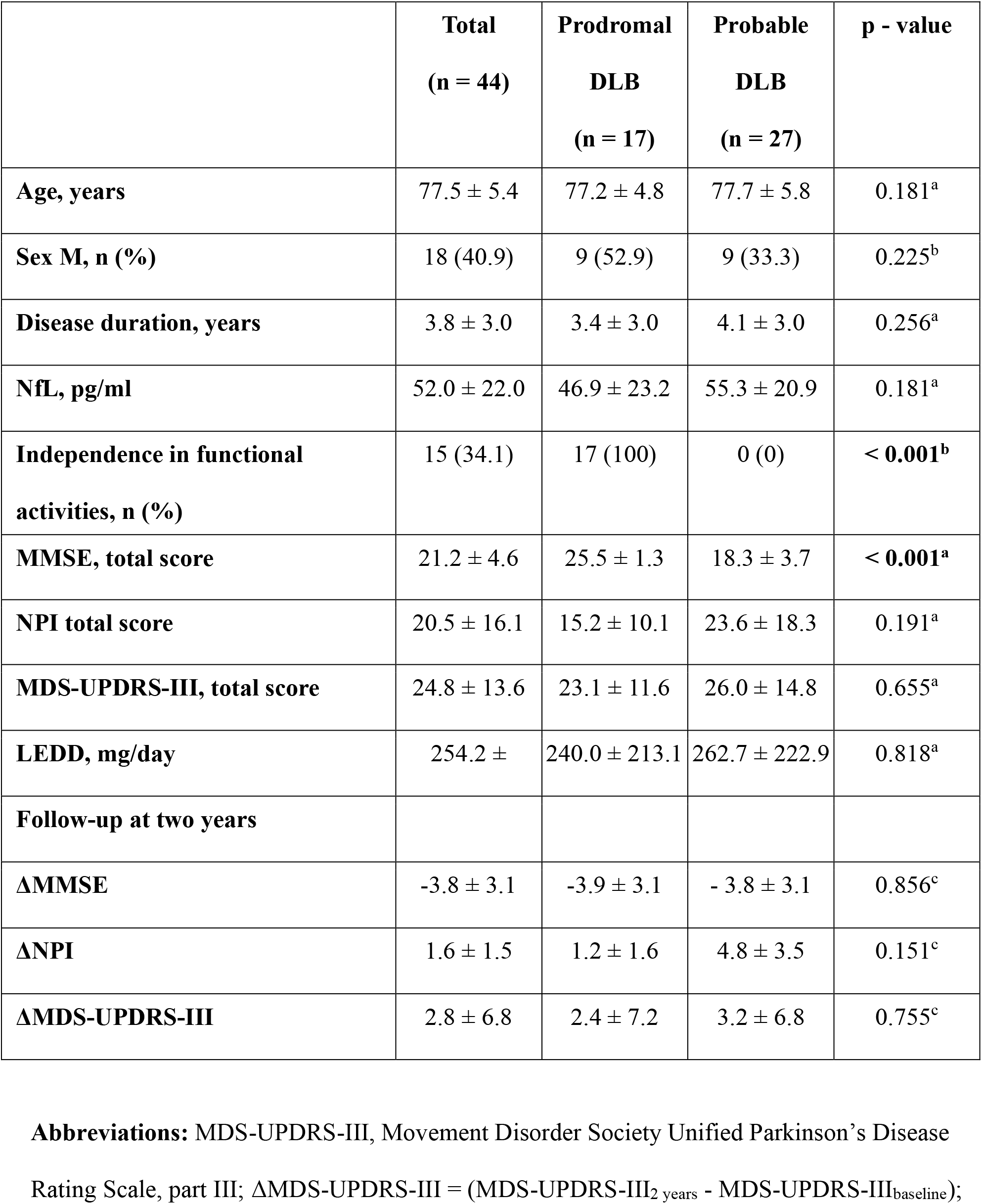
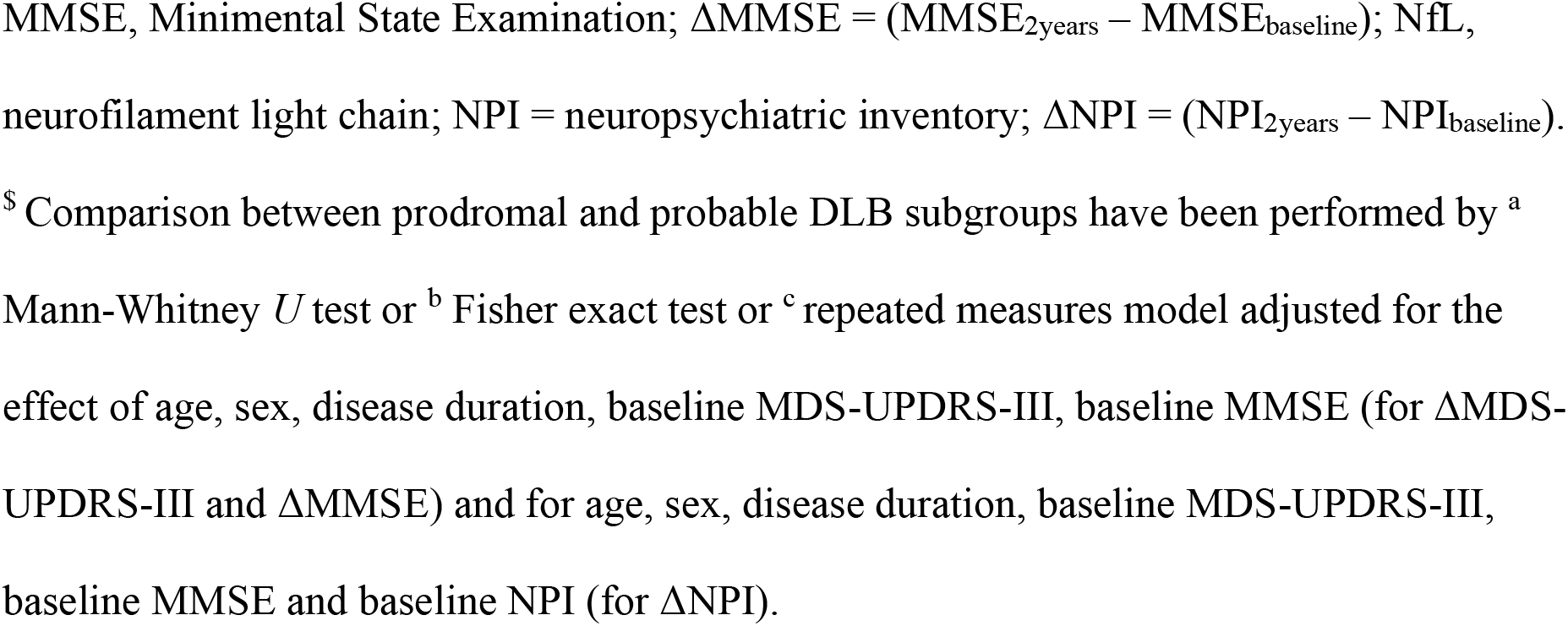
Clinical and demographic characteristics of prodromal DLB and probable DLB patients at baseline and after 2-years follow-up.

|  | **Total**<br>**(n = 44)** | **Prodromal**<br>**DLB**<br>**(n = 17)** | **Probable**<br>**DLB**<br>**(n = 27)** | **p - value** |
| --- | --- | --- | --- | --- |
| **Age, years** | 77.5 ± 5.4 | 77.2 ± 4.8 | 77.7 ± 5.8 | 0.181<sup>a</sup> |
| **Sex M, n (%)** | 18 (40.9) | 9 (52.9) | 9 (33.3) | 0.225<sup>b</sup> |
| **Disease duration, years** | 3.8 ± 3.0 | 3.4 ± 3.0 | 4.1 ± 3.0 | 0.256<sup>a</sup> |
| **NfL, pg/ml** | 52.0 ± 22.0 | 46.9 ± 23.2 | 55.3 ± 20.9 | 0.181<sup>a</sup> |
| **Independence in functional activities, n (%)** | 15 (34.1) | 17 (100) | 0 (0) | **< 0.001<sup>b</sup>** |
| **MMSE, total score** | 21.2 ± 4.6 | 25.5 ± 1.3 | 18.3 ± 3.7 | **< 0.001<sup>a</sup>** |
| **NPI total score** | 20.5 ± 16.1 | 15.2 ± 10.1 | 23.6 ± 18.3 | 0.191<sup>a</sup> |
| **MDS-UPDRS-III, total score** | 24.8 ± 13.6 | 23.1 ± 11.6 | 26.0 ± 14.8 | 0.655<sup>a</sup> |
| **LEDD, mg/day** | 254.2 ± | 240.0 ± 213.1 | 262.7 ± 222.9 | 0.818<sup>a</sup> |
| **Follow-up at two years** |  |  |  |  |
| **ΔMMSE** | -3.8 ± 3.1 | -3.9 ± 3.1 | - 3.8 ± 3.1 | 0.856<sup>c</sup> |
| **ΔNPI** | 1.6 ± 1.5 | 1.2 ± 1.6 | 4.8 ± 3.5 | 0.151<sup>c</sup> |
| **ΔMDS-UPDRS-III** | 2.8 ± 6.8 | 2.4 ± 7.2 | 3.2 ± 6.8 | 0.755<sup>c</sup> |

**Abbreviations:** MDS-UPDRS-III, Movement Disorder Society Unified Parkinson's Disease
Rating Scale, part III; ΔMDS-UPDRS-III = (MDS-UPDRS-III<sub>2 years</sub> - MDS-UPDRS-III<sub>baseline</sub>);
MMSE, Minimental State Examination; $\Delta\text{MMSE} = (\text{MMSE}_{2\text{years}} - \text{MMSE}_{\text{baseline}})$ ; NfL, neurofilament light chain; NPI = neuropsychiatric inventory; $\Delta\text{NPI} = (\text{NPI}_{2\text{years}} - \text{NPI}_{\text{baseline}})$ .
<sup>§</sup> Comparison between prodromal and probable DLB subgroups have been performed by <sup>a</sup> Mann-Whitney *U* test or <sup>b</sup> Fisher exact test or <sup>c</sup> repeated measures model adjusted for the effect of age, sex, disease duration, baseline MDS-UPDRS-III, baseline MMSE (for $\Delta\text{MDS-UPDRS-III}$ and $\Delta\text{MMSE}$ ) and for age, sex, disease duration, baseline MDS-UPDRS-III, baseline MMSE and baseline NPI (for $\Delta\text{NPI}$ ).

**Figure 1.**
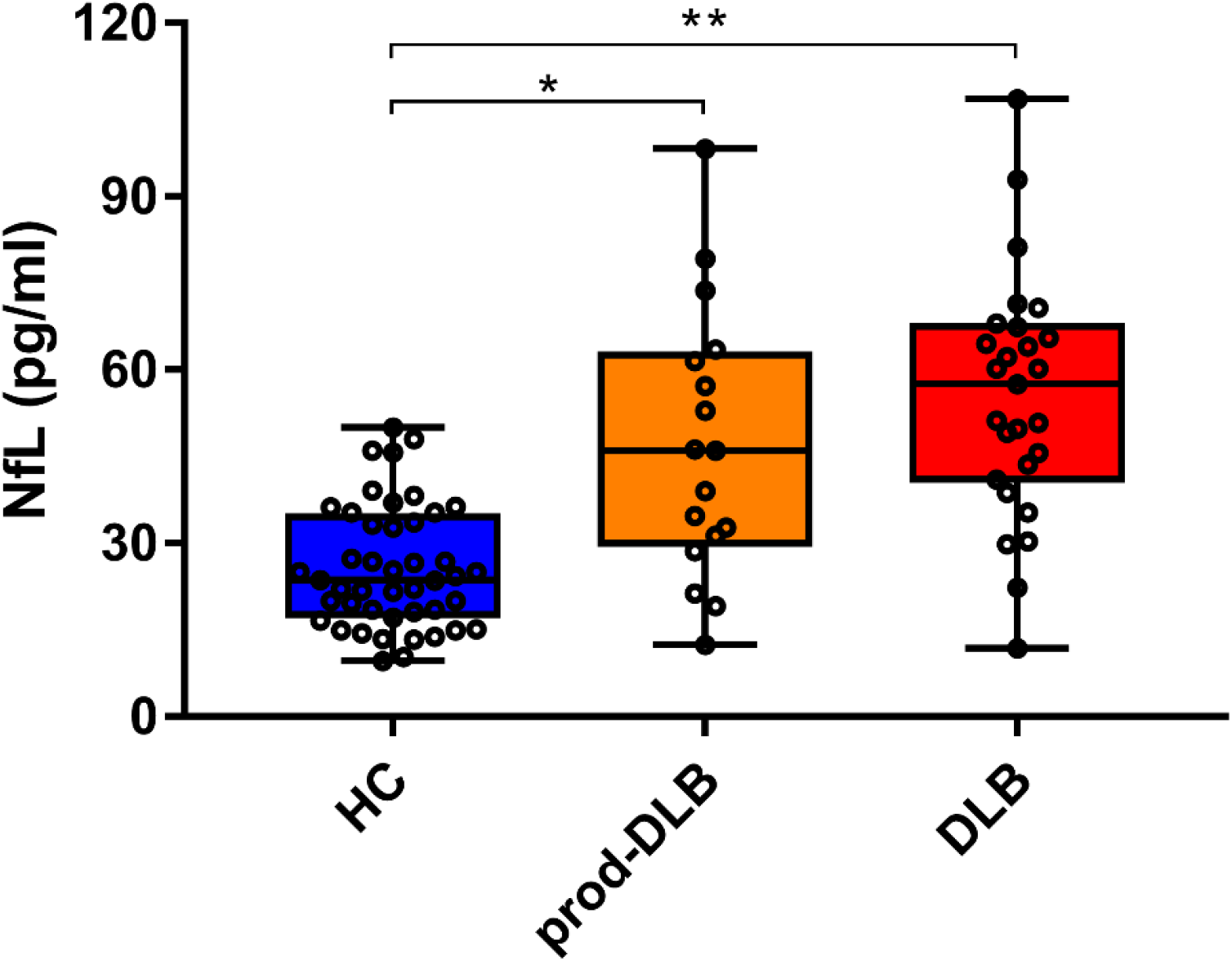
Box and whiskers plot showing plasma NfL levels in HC, prod-DLB and DLB patients. Data are shown as medians with 25°–75° percentiles (boxes), min-max values (whiskers) and individual values (dots). DLB = probable Dementia with Lewy Bodies; HC = healthy controls; NfL = neurofilament light chain; prod-DLB = prodromal Dementia with Lewy Bodies; * p = 0.001; ** p < 0.001.

In total DLB cohort, plasma NfL correlated with age (r = 0.369; p = 0.014) and exhibited similar distribution in male and female subjects. At the time of baseline assessment, no correlation between NfL levels and disease duration or severity of motor, cognitive and neuropsychiatric features was detected in analyses adjusted for age and sex (in the whole group and separately for prodromal and clinical cases).

### Plasma NfL as predictor of progression

At follow up, the total group of DLB patients exhibited a mean decrease of 3.8 ± 3.1 points at MMSE score, a mean increase of 1.6 ± 1.5 points in total NPI and 2.8 ± 6.8 points in MDS-UPDRS-III score. In the model without NfL (model 1, supplementary Table 1), no predictors of changes in MMSE were identified (R^2^ = 0.160) while the inclusion of NfL (model 2) increased the prediction of the linear model (R^2^ = 0.356) and age (p = 0.047) and NfL (p = 0.034) were the only significant predictors of changes in MMSE (Table 2). No predictors of NPI progression were detected in linear regression model, whereas male gender was the only predictor of ΔMDS-UPDRS-III in the model with (β = 0.387, p = 0.023) and without (β = 0.373, p = 0.026) the inclusion of NfL.

**Table 2.**
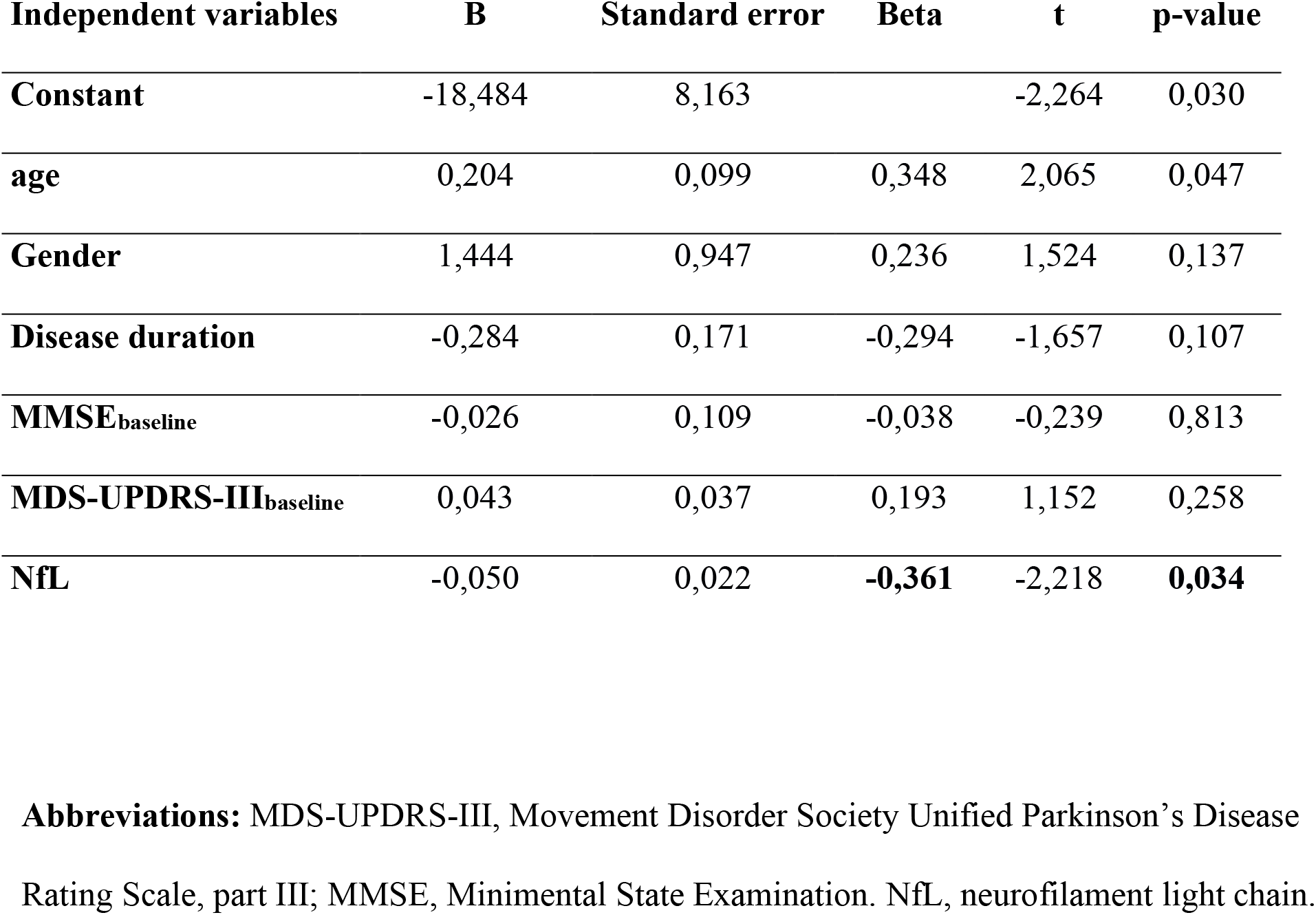
Multivariable linear regression model for cognitive progression in total DLB cohort defined by changes in MMSE score including demographic, clinical baseline variables and plasma NfL levels.

At follow-up, 10 out of 17 prodromal cases converted to dementia with Lewy bodies; converters did not differ for demographic or clinical baseline variables and exhibited slightly higher but not significant NfL levels compared with non-converters (51.4 ± 22.9 vs 40.5 ± 23.9; p = 0.37).

## DISCUSSION

This is the first longitudinal study addressing plasma NfL as an early diagnostic and prognostic marker in DLB patients. We found higher NfL levels in DLB patients compared with controls from the prodromal stages and we demonstrated for the first time that baseline NfL levels predicted cognitive decline over a follow-up of 2 years.

The study included consecutive patients with a clinical diagnosis of probable DLB and a subset of prodromal cases who underwent an extensive motor, cognitive and behavioral assessment. We demonstrated that clinical DLB cases exhibited increased plasma NfL levels compared with controls, in line with several cross-sectional studies focused on neurodegenerative disorders [7,19]. Furthermore, the findings showed that cases who retrospectively fulfilled the newly proposed prodromal DLB criteria [3] exhibited already increased plasma NfL levels compared with controls. These important results strongly support the usefulness of plasma NfL as diagnostic marker of DLB already in the prodromal stages, in line with the work of Delaby and colleagues [21] based on CSF data on DLB with and without dementia.

In addition, our study aimed to define whether plasma NfL levels might be of prognostic value, a still unsolved issue with great relevance for the upcoming disease-modifying trials. Indeed, the identification of consistent disease progression markers is still an open challenge for DLB. Recently, a growing number of studies identified EEG, imaging or CSF abnormalities to be associated with faster progression already in early DLB stages [22–24]. However, these techniques definitively need further validations to be applied in clinical practice.

In the present study, we identified higher blood NfL levels as a significant predictor of faster cognitive progression in DLB patients at 2 year follow-up. These results were strengthened by a linear regression model taking into account age, sex and the baseline clinical data for both clinical and prodromal DLB patients. These findings are at variance with two earlier studies performed on CSF, that failed to demonstrate a relationship between NfL levels and annual MMSE decline [25][26] in smaller group of DLB patients. Of note, the prognostic value of NfL was specifically demonstrated for cognitive changes, whereas was not confirmed for behavioral and motor progression in the cohort. This fit with data coming from studies on Alzheimer’s disease [12] but also with Parkinson’s disease in which higher NfL levels have been more consistently associated with cognitive decline [10].

Several limitations should be acknowledged. First, a 2-year follow-up period is a short interval to consider these findings as definitive, and further validations in longer and larger longitudinal multicentre studies are needed in order to evaluate the long-term value of NfL in predicting progression on disability milestones. Second, the small sample size required the use of non-parametric statistical analysis and limited the number of covariates in linear regression model, and since this was an exploratory study we did not adjust for multiple statistical comparisons. Third, the study evaluated clinical outcomes and the findings need to be replicated against imaging, CSF of neurophysiological or other promising peripheral markers, such as P-Tau [12,27]. Diagnosis was clinical associated with DAT imaging but without autopsy confirmation, and the prodromal criteria were applied retrospectively.

However, the longitudinal design and standardized comprehensive clinical assessments and use of the most recent consensus diagnostic criteria ensures a high diagnostic accuracy.

Limitations notwithstanding, this is the first study demonstrating the diagnostic and prognostic value of plasma NfL in DLB already from prodromal stages.

## Data Availability

Data are available upon reasonable request to the corresponding Author.

## ACKNOWLEDGMENTS

The authors thanks all the participants for their participation.

Study funding: The recruitment of patients was partially supported by BIOMANE Project from the Health and Wealth research grants 2016 of the University of Brescia [grant number NP 1471, DMA].

## Relevant conflicts of interest/financial disclosures (for the past two years)

all the authors report no disclosures related to this manuscript. Other disclosures are reported below.

Andrea Pilotto served in the advisory board of Z-cube (technology division of Zambon pharmaceuticals); he received honoraria from Z-cube s.r.l., Biomarin, Zambon, Nutricia and Chiesi pharmaceuticals. He received research support from Vitaflo Germany and Zambon Italy.

Alberto Imarisio has no financial conflicts to disclose.

Claudia Carrarini has no financial conflicts to disclose.

Mirella Russo has no financial conflicts to disclose.

Stefano Masciocchi has no financial conflicts to disclose.

Stefano Gipponi has no financial conflicts to disclose.

Elisabetta Cottini has no financial conflicts to disclose.

Dag Aarsland has received research support and/or honoraria from Astra-Zeneca, H.

Lundbeck, Novartis Pharmaceuticals, Evonik, and GE Health, and served as paid consultant for H. Lundbeck, Eisai, Heptares, Mentis Cura, Eli Lilly and Biogen.

Henrik Zetterberg has served at scientific advisory boards for Denali, Roche Diagnostics, Wave, Samumed, Siemens Healthineers, Pinteon Therapeutics and CogRx, has given lectures in symposia sponsored by Fujirebio, Alzecure and Biogen, and is a co-founder of Brain Biomarker Solutions in Gothenburg AB (BBS), which is a part of the GU Ventures Incubator Program (outside submitted work).

Nicholas Ashton has no financial conflicts to disclose.

Abdul Hye has no financial conflicts to disclose.

Laura Bonanni received unrelated national grants from the Italian Ministry of Health, from European Commission (PD MIND project, IMI call) and from Mentis cura, Oslo srl; fees from G.E. Healthcare for teaching courses. Served as reviewer for NIHhas no financial conflicts to disclose.

Alessandro Padovani is consultant and served on the scientific advisory board of GE Healthcare, Eli-Lilly and Actelion Ltd Pharmaceuticals, received speaker honoraria from Nutricia, PIAM, Lansgstone Technology, GE Healthcare, Lilly, UCB Pharma and Chiesi Pharmaceuticals. He is founded by Grant of Ministry of University (MURST).

---

**Supplementary Table 1.**
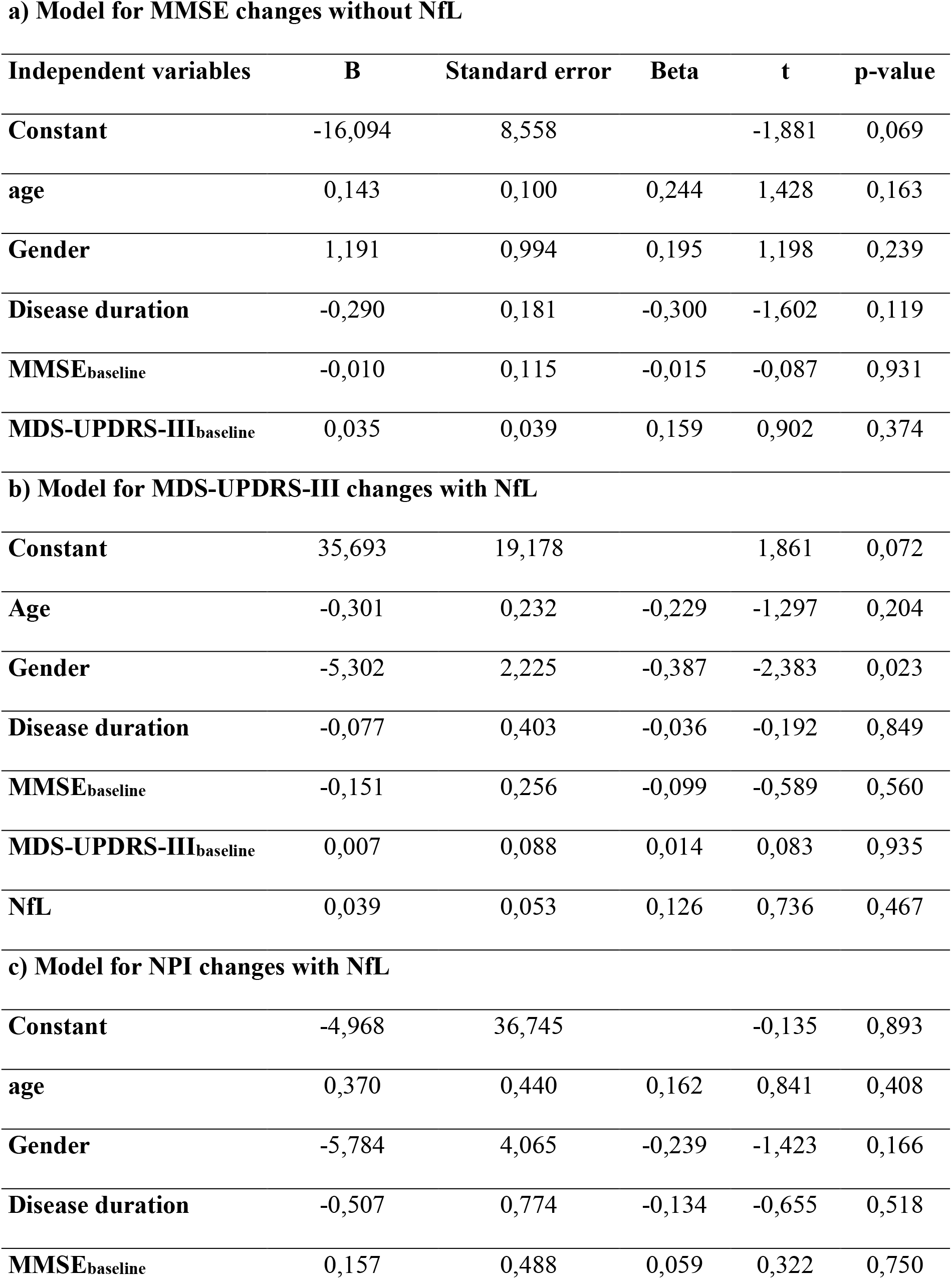
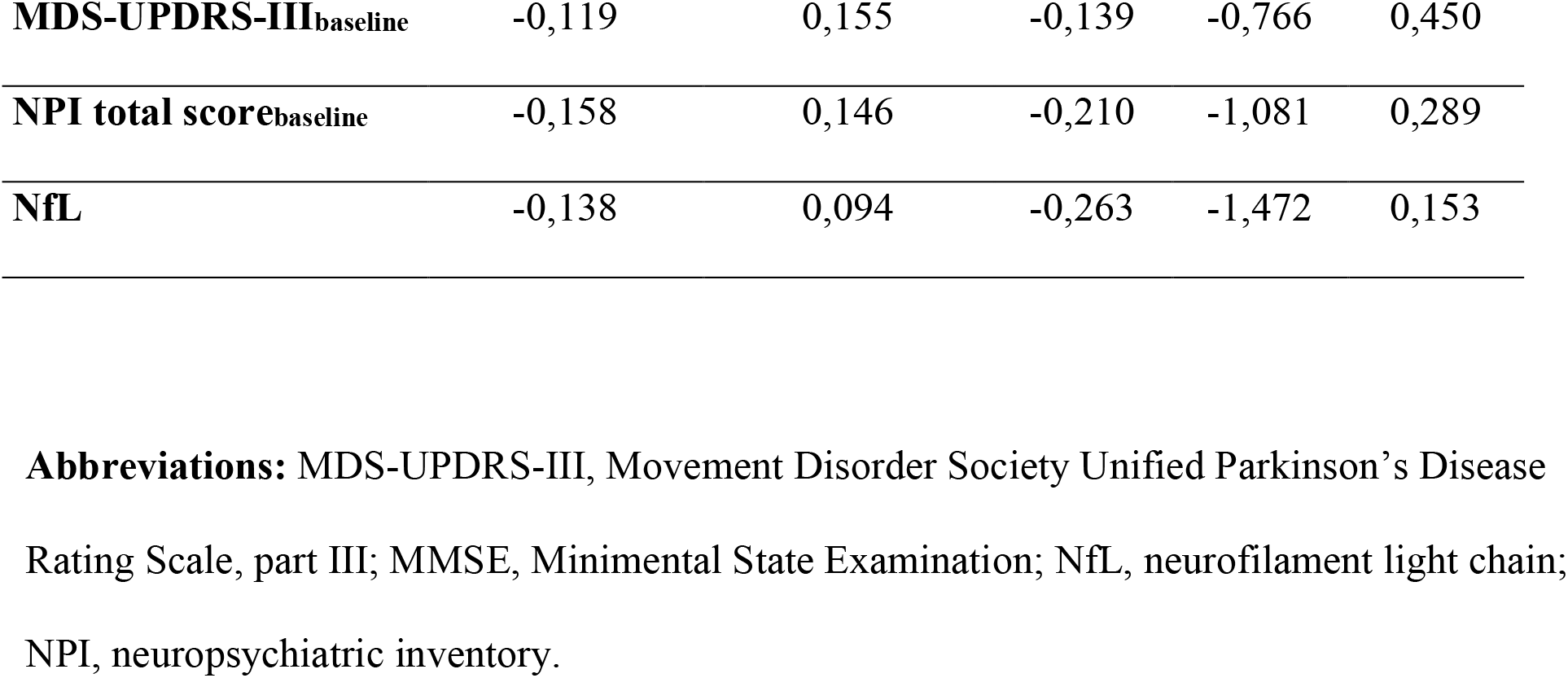
Multivariable linear regression for factors associated with cognitive, motor and behavioral progression. Model a) included demographic and clinical variables but no NfL at baseline for cognitive progression in total DLB cohort defined by changes within MMSE score. Models b) and c) showed the predicting models of motor progression defined by MDS-UPDRS-III (b) and behavioral abnormalities defined by NPI changes (c)

**a) Model for MMSE changes without NfL**
| **Independent variables** | **B** | **Standard error** | **Beta** | **t** | **p-value** |
| --- | --- | --- | --- | --- | --- |
| **Constant** | -16,094 | 8,558 |  | -1,881 | 0,069 |
| **age** | 0,143 | 0,100 | 0,244 | 1,428 | 0,163 |
| **Gender** | 1,191 | 0,994 | 0,195 | 1,198 | 0,239 |
| **Disease duration** | -0,290 | 0,181 | -0,300 | -1,602 | 0,119 |
| **MMSE<sub>baseline</sub>** | -0,010 | 0,115 | -0,015 | -0,087 | 0,931 |
| **MDS-UPDRS-III<sub>baseline</sub>** | 0,035 | 0,039 | 0,159 | 0,902 | 0,374 |
| **b) Model for MDS-UPDRS-III changes with NfL** |  |  |  |  |  |
| **Constant** | 35,693 | 19,178 |  | 1,861 | 0,072 |
| **Age** | -0,301 | 0,232 | -0,229 | -1,297 | 0,204 |
| **Gender** | -5,302 | 2,225 | -0,387 | -2,383 | 0,023 |
| **Disease duration** | -0,077 | 0,403 | -0,036 | -0,192 | 0,849 |
| **MMSE<sub>baseline</sub>** | -0,151 | 0,256 | -0,099 | -0,589 | 0,560 |
| **MDS-UPDRS-III<sub>baseline</sub>** | 0,007 | 0,088 | 0,014 | 0,083 | 0,935 |
| **NfL** | 0,039 | 0,053 | 0,126 | 0,736 | 0,467 |
| **c) Model for NPI changes with NfL** |  |  |  |  |  |
| **Constant** | -4,968 | 36,745 |  | -0,135 | 0,893 |
| **age** | 0,370 | 0,440 | 0,162 | 0,841 | 0,408 |
| **Gender** | -5,784 | 4,065 | -0,239 | -1,423 | 0,166 |
| **Disease duration** | -0,507 | 0,774 | -0,134 | -0,655 | 0,518 |
| **MMSE<sub>baseline</sub>** | 0,157 | 0,488 | 0,059 | 0,322 | 0,750 |
| **MDS-UPDRS-III**<sub>baseline</sub> | -0,119 | 0,155 | -0,139 | -0,766 | 0,450 |
| **NPI total score**<sub>baseline</sub> | -0,158 | 0,146 | -0,210 | -1,081 | 0,289 |
| **NfL** | -0,138 | 0,094 | -0,263 | -1,472 | 0,153 |

**Abbreviations:** MDS-UPDRS-III, Movement Disorder Society Unified Parkinson's Disease Rating Scale, part III; MMSE, Minimental State Examination; NfL, neurofilament light chain; NPI, neuropsychiatric inventory.

